# Promotores de Salud: Feasibility of a pilot community health worker program to reach and engage Hispanic and Latino people with Parkinson’s disease

**DOI:** 10.1101/2025.05.02.25326745

**Authors:** Evelyn Stevens, M. Felice Ghilardi, Alessandro Di Rocco, Maria Lima, Elisa Tatti, Donna Sperlakis, Jennifer Fearday, Meri-Margaret Deoudes, Nora Vanegas-Arroyave, Denise Hernandez, Paulina Latapi-Gonzalez, Christiana Evers

## Abstract

Parkinson’s disease (PD), a progressive neurodegenerative disorder, manifests with motor and non-motor symptoms. Despite similar incidence, clinical care for Hispanic patients with PD is poor compared to white non-Hispanic individuals. As a result, their participation in PD research also does not reflect expected values. These disparities suggest an underutilization of quality healthcare, socio-economic disadvantage, stigma, and cultural differences. Using a community health worker pilot program, we trained 298 *Promotores de Salud* to reach, educate, and engage the Hispanic community in healthcare. Outcomes demonstrated improved knowledge of PD among Promotores, as well as increased access and utilization of educational resources.

---

Parkinson’s disease (PD), the world’s fastest-growing neurodegenerative disease, is characterized by motor and non-motor manifestations. Although the incidence of PD in the United States appears to be similar amongst races and ethnicities,^1^ there is strong evidence of inequities in PD care, with the best predictors for optimal care being high socio-economic level, marital status, urban living, and being white.^2^ Within the Hispanic community (defined as Spanish-speaking people of Central and South American origin), diagnosis and management of PD lags behind that of non-Hispanic whites. Consequently, the participation of Hispanic patients with PD in research is also low, with participation rates varying from 1.6 to 5.4%, ^3,4^ despite Hispanics representing at least 18% of the US population.^1^ This discrepancy may reflect limited healthcare access or underutilization of healthcare services, low health literacy, socio-economic disadvantages, stigma, and differing cultural perceptions of PD manifestations.^5^ This critical gap needs to be addressed to verify the generalization of research and to make sure that Hispanic patients with PD receive appropriate care.

Community health workers (CHWs), commonly referred to as *health educators, peer advocates*, and among the Hispanic and Latino community, *promotores de salud*, have made significant impacts engaging underrepresented communities in care and research by connecting individuals and families to resources and support.^6^ CHWs are frontline healthcare workers who share a common understanding or experience, often cultural and linguistic, with the community they serve, enabling them to develop trusting relationships within communities.^7^ Existing literature suggests the efficacy of CHWs in improving health outcomes across several diseases and conditions, including cancer,^8– 10^ diabetes,^11–13^ mental health;^14^ and within the Hispanic and Latino community.^15–18^

There is a paucity of research on CHWs reaching individuals living with neurological conditions and even fewer studies on CHWs reaching Hispanic and Latinos living with neurological conditions. Studies to date have been focused on Alzheimer’s disease and dementia ^19,20^, and to our knowledge, there are no published studies on CHW programs aimed at reaching Hispanics and Latinos living with PD. Expanding on the success of *Promotores de Salud* programs across other conditions and to address the overall gaps with programs tailored and targeted for the Hispanic and Latino community living with PD, we piloted the feasibility of a community health worker training program aimed at improving knowledge of PD and access to resources among established CHWs that can return to their communities and provide education to Hispanics and Latinos living with PD.

## Methods

The Parkinson’s Foundation established the *Promotores de Salud (Promotores)* training program through partnerships with community-based organizations and CHWs already embedded in the Hispanic community. As with other CHW models, the *Promotores* program is based on the concept that trusted individuals within communities can be the best outreach and education conduits. Seven locations were identified as target markets based on established CHW networks; incidence, prevalence, and healthcare utilization of those living with PD enrolled in Medicare in 2019; ^21^ and census data of Hispanic/Latino populations. These locations include: Arlington, Texas; El Paso, Texas; Houston, Texas; Yuma, Arizona; Springfield, Illinois; and Chicago, Illinois. CHWs were invited to participate in the program through their affiliated community health worker group and/or organizations. This approach maximized sustainability by identifying CHWs who would be committed to reaching the Hispanic community and garnering support from CHW groups/organizations with curriculum development and data collection.

Guided by best practices in patient engagement,^22^ the training workshops were developed with input from the Parkinson’s community (people with lived experience, PD specialists) and a community health worker organization that reviewed and approved the training for core competencies.^23^ Content included early signs and causes of PD, motor and non-motor symptoms, treatments, and how to access available resources and support from the Parkinson’s Foundation *(Table 1)*. The workshops were led by Spanish-speaking movement disorder specialists and Parkinson’s Foundation staff. Pre and post-test surveys were provided in Spanish for participants to complete before and after the training, respectively. Given that this is the first study we are aware of aimed at developing a *Promotores de Salud* program for the PD community, the surveys included self-rated familiarity with PD (one item), knowledge of PD, and where to find resources on PD (four items). The program was reviewed by Advarra Institutional Review Board and determined to be exempt research. Descriptive statistics were used for pre and post-test analysis. Shipping of our Spanish resources was tracked over time to assess impact of the *Promotores* program and the information needs of the Hispanic and Latino community.

**Table 1.** Example of a Schedule & Agenda *“Parkinson’s Disease 101: How to Talk About Parkinson’s in Your Community”*.

|  |  |
| --- | --- |
| 9:30 am (30 min) | Sign in, breakfast, networking, Pretest |
| 10:00 am (10 min) | Parkinson’s Foundation Introduction |
| 10:10 am (60 min) | Parkinson’s Disease: Introduction, Causes, factors/Activity 1;<br>Symptoms/Activity 2 |
| 11:10 am (10 min) | BREAK |
| 11:20 am (30 min) | How to manage PD and what resources are available? |
| 11:50 am (20 min) | Q&A |
| 12:10 am (30 min) | Activity 3- resources |
| 12:40 am (20 min) | Report Out |
| 1:00 pm (5 min) | Conclusions |
| 1:10 am (20 min) | Posttest/Evaluation/Networking;<br>Certificate upon completion |

## Results

The Parkinson’s Foundation provided seven training sessions to 298 CHWs from November 2022 through December 2023. The most endorsed affiliations by CHWs included community-based organizations (49%), hospitals (5%), local health jurisdictions (5%), and schools or universities (5%). When asked about their connection to PD, most (62%) indicated having had no connection to PD, and 26% indicated having personal connections to PD (e.g., they were people with PD or were friend, family, parent, caregiver, or spouse of a person with PD).

Of the 298 CHWs trained, 152 completed both pre- and post-test surveys. Familiarity with PD and management increased following the training, with 96 (63%) indicating no familiarity before the training while afterward, 148 (98%) indicating knowing some basic facts or being very familiar with PD and management. Similarly, understanding non-movement symptoms and risk factors of PD improved after the training *(Table 2)*. Notably, the percentage of CHWs that indicated knowing where to find resources about PD rose from 20% (n=30) before the training to 97% (n=147) afterward.

**Table 2.** Promotores de Salud Training – Pretest and Posttest Survey (n=152)

| Item | Pretest (n, %) | Posttest (n, %) |
| --- | --- | --- |
| Please select the answer that best describes your knowledge about Parkinson's Disease (PD) |  |  |
| I am not familiar with PD | 96 (63%) | 3 (2%) |
| I know some basic facts about PD | 49 (32%) | 85 (56%) |
| I am very familiar and have great knowledge of PD and care | 5 (3%) | 64 (42%) |
| No response | 3 (2%) | 0 |
| Does PD cause symptoms other than movement symptoms? |  |  |
| Yes | 52 (34%) | 141 (93%) |
| No | 6 (4%) | 2 (1%) |
| I don't know | 90 (59%) | 5 (3%) |
| No response | 5 (3%) | 5 (3%) |
| What causes PD? |  |  |
| Environmental factors | 2 (1%) | 0 |
| Genetics | 18 (12%) | 3 (2%) |
| Advanced age | 11 (7%) | 2 (1%) |
| All of the above | 93 (61%) | 146 (96%) |
| I don't know | 27 (18%) | 0 |
| I know where to find resources and education about PD? |  |  |
| Yes | 29 (19%) | 147 (97%) |
| No | 27 (18%) | 2 (1%) |
| I don't know | 94 (62%) | 2 (1%) |
| No response | 2 (1%) | 2 (1%) |

Parkinson’s Foundation Spanish educational resources were tracked during the trainings. There were 19,857 publications shipped during the *Promotores* trainings, ranging from 358 in November 2022 to 1,474 in December 2023 (average per month: 1,416). The most requested resources were fact sheets and the top two were: About Parkinson’s Disease and Exercise Recommendations.

## Discussion

Growing evidence indicates that Hispanic and Latino people with PD are at an increased risk of poor health outcomes, lower quality care, and low participation in research.^5, 22–24^ To address these disparities, we developed and piloted a *Promotores de Salud* program to train CHWs to reach, educate, and engage the Hispanic and Latino community with PD. This program is among the first to equip CHWs with PD-specific knowledge.

Findings indicated the feasibility of training CHWs on clinical aspects and importance of engaging the Hispanic community in healthcare. Most of the CHWs had no familiarity with PD before this training, particularly non-motor symptoms, underscoring the importance of these targeted educational efforts. Additionally, CHWs returned to their communities understanding where to find resources and education for community members. Throughout the trainings, the most ordered resource were our Spanish fact sheets, aligning with the health literacy needs of this community,^27^ concise, easy to understand health information, with visuals, resonates well with the Hispanic and Latino community. The fact sheets “About Parkinson’s” and “Exercise Recommendations” were the most popular. Exercise is a well-studied component of PD management,^28^ and the strong demand for exercise-focused information suggests that this program may also be effective in indirectly improving low rates of physical activity among Latino adults.^29^

The *Promotores de Salud* training and findings are subject to limitations. First, only about half of the CHWs who were trained completed both the pretest and posttest surveys, potentially limiting the generalizability of the results Second, tracking the orders of our Spanish-language resources was the only measure of impact. Other factors such as the Parkinson’s Awareness Month and National Hispanic Heritage Month campaigns may have influenced resource requests during this period. Evaluating the extent of CHWs outreach would have provided a more comprehensive assessment of the program’s real-world impact. Future research could measure the impact by including a research component in the training and tracking of the reach of the CHWs and the number of Hispanic and Latino people who enroll in research studies. This may improve the representation of the Hispanic and Latino communities in clinical trials or other types of research studies that advance our knowledge of PD,^30^ and enhances our ability to design future trials that include Hispanic and Latino people with PD.^31^ Despite these limitations, this *Promotores de Salud* program provided CHWs with PD education in their native language and access to PD resources, equipping them with tools to raise awareness in their communities. These efforts begin to fill a critical gap in the current literature and emphasize the need for researchers, healthcare professionals, and community-based organizations to continue thinking about the importance of CHW interventions for Hispanic and Latino people with PD.

## Data Availability

All data produced in the present work are contained in the manuscript

## Acknowledgments

The authors gratefully acknowledge the contributions of people with Parkinson’s, community health workers, Dr. Katherline Longardner, Dr. Juan Ramirez-Castaneda, and Dr. Charenya Anandan to developing this research engagement model.

